# Detecting adrenal lesions on 3D CT scans using a 2.5D deep learning model

**DOI:** 10.1101/2023.02.22.23286184

**Authors:** Sanson T. S. Poon, Fahmy W. F. Hanna, François Lemarchand, Cherian George, Alexander Clark, Simon Lea, Charlie Coleman, Giuseppe Sollazzo

## Abstract

Many cases of adrenal lesions, known as adrenal incidentalomas, are discovered incidentally on CT scans performed for other medical conditions. Whilst they are largely benign, these lesions can be secretory and/or malignant. Therefore, early investigation is crucial to promptly and efficiently manage those requiring intervention whilst to reassuring the remaining majority in a timely manner. Traditionally, the detection of adrenal lesions on CT scans relies on manual analysis by radiologists, which can be time-consuming and unsystematic. Using AI and deep learning, we examined whether or not applying these technology can augment the detection of adrenal incidentalomas in CT scans. We developed a 2.5D deep learning model to perform image classification on 3D CT scans of patients to classify between lesion and healthy adrenal glands. When tested on an independent test set, our 2.5D model obtained an AUC of the ROC curve of 0.95, and a classification sensitivity of 0.86, and specificity of 0.89. These results suggest that deep learning may be a promising tool for detecting adrenal lesions and improving patient care.

## 1 Introduction

Autopsy studies reveal a statistic that as many as 6% of all natural deaths displayed a previously undiagnosed adrenal lesion (e.g., Kloos et al., 1995; Mansmann et al., 2004; Anagnostis et al., 2009). Such lesions are also found incidentally (and are therefore referred to as adrenal incidentalomas) in approximately 1% of chest or abdominal computed tomography (CT) studies (e.g., Abecassls et al., 1985; Bitter and Ross, 1989; Caplan et al., 1994; Song et al., 2008; Sherlock et al., 2020). It is estimated that these lesions could affect up to 50,000 patients annually in the United Kingdom (UK), with significant impact on patient health, including 10 to 15% cases of excess hormone production, together with 1 to 5% cases of cancer (Hanna et al., 2020). It is a significant challenge to the health care system, in a standardised way, to promptly reassure the majority with no abnormalities whilst effectively and promptly focusing on those with hormone excess or cancers. Issues include: over-reporting (false positives), causing patient anxiety and unnecessary investigations (wasting resources of the health care system); and under-reporting (missed cases), with potentially fatal outcomes. This has major impacts on patient well-being and clinical/cost-effectiveness.

The main aim of this study is to examine whether or not using Artificial Intelligence (AI) can improve the detection of adrenal incidentalomas in CT scans. Previous studies have suggested that AI has the potential in distinguishing different types of adrenal lesions (e.g., Yi et al., 2018; Elmohr et al., 2019; Moawad et al., 2021; Kusunoki et al., 2022). In this study, we specifically focused on detecting the presence of any type of adrenal lesion in CT scans. To demonstrate this proof-of-concept, we investigated the potential of applying deep learning techniques to predict the likelihood of a CT abdominal scan presenting as ‘normal’ or ‘abnormal’, the latter implying an adrenal lesion.

Recently, applying a standard 3D deep learning model for classification on medical CT scans has shown some promising results (e.g., Nie et al., 2016; Kruthika et al., 2019; Zhou et al., 2019; Singh et al., 2020). However, the acquisition of a sufficient amount of CT scans for 3D deep learning models training is challenging (e.g., a high operating cost, limited number of available CT scanners, and patients exposure to radiation). In many cases, the performance of 3D deep learning models are limited by the small and non-diversified dataset. Training, validating, and testing the model with a small dataset can lead to many disadvantages, for example, a high risk of overfitting the training-validation set (low prediction ability on an unseen test set), and evaluating the model performance within the ambit of small number statistics (e.g., an unrepresentative test set can result in the test accuracy much lower/higher than the underlying model performance).

With a small number of CT scans available in our dataset, applying a traditional 3D deep learning model might not be the best approach due to the high dimensional complexity of the input images. Instead, we applied a 2.5D deep learning model approach in this proof-of-concept study (Section 3). A CT scan is a 3D image composed of a number of 2D slices but it can also be transformed into multiple so-called 2.5D images (see Section 3.1). A 2.5D image contains a small subset of adjacent 2D slices. It has the same pixel height and width as a 2D image but can also retain some 3D features from the original CT scan. Applying a 2.5D model allows the deep learning model to learn from 3D features while increasing the number of training and testing data points in this study by ≳ 43 times.

This paper is structured as follows. In Section 2, we discuss our dataset for this study and the data pre-processing steps for this study. In Section 3, we describe our 2.5D deep learning model for adrenal lesion classification. In Section 4, we present the performance and results of our adrenal lesion classification model. Finally, we discuss our results and draw conclusions in Section 5.

## 2 Materials and dataset preprocessing

All CT scan examples included in this study were provided by the University Hospitals of North Midlands NHS Trust. The entire dataset is completely anonymised and does not contains personal information from patients, such as the patients’ age, gender, or ethnicity.

### 2.1 The dataset

The dataset contained a study of 100 anonymised patients from the University Hospitals of North Midlands NHS Trust. 50 of these patients did not have any sign of adrenal lesions (labelled as ‘normal’ patients), and the other 50 patients had signs of different types of adrenal lesions (e.g., adenomas or carcinomas; labelled as ‘abnormal’ patients). For abnormal cases, the dataset did not contain information on the location (left or right adrenal gland) of the lesion. In total, across the 100 patients, there are 234 3D CT scans (151 normal and 83 abnormal) considered in this study (Table 1).

**Table 1:** A summary of the number of patients and scans contained in the dataset for this study.

|  | Normal | Abnormal |
| --- | --- | --- |
| No. of patients | 50 | 50 |
| No. of 3D CT scans | 151 | 83 |
|  | Training | Test |
| No. of patients | 80 | 20 |
| No. of 3D CT scans | 194 | 40 |
| No. of 2D axial slices | 8340 | 1863 |

All the CT scans included in this study fulfilled some general selection criteria. Firstly, we only selected the CT scans that covered the entire adrenal glands of the patients (both left and right adrenal glands) in all three dimensions. All the scans have a slice thickness between 0.5 and 3.0 mm, and with the same iodinated contrast density.

### 2.2 Region of interest

To focus on our region of interest (ROI) of this study, which is the adrenal glands of the patients, we manually cropped the CT scans to exclude some irrelevant body parts. The cropping applied to all three dimensions, including a 1D cropping to select the appropriate axial slices and a 2D cropping on the axial slices (Figure 1).

**Figure 1:**
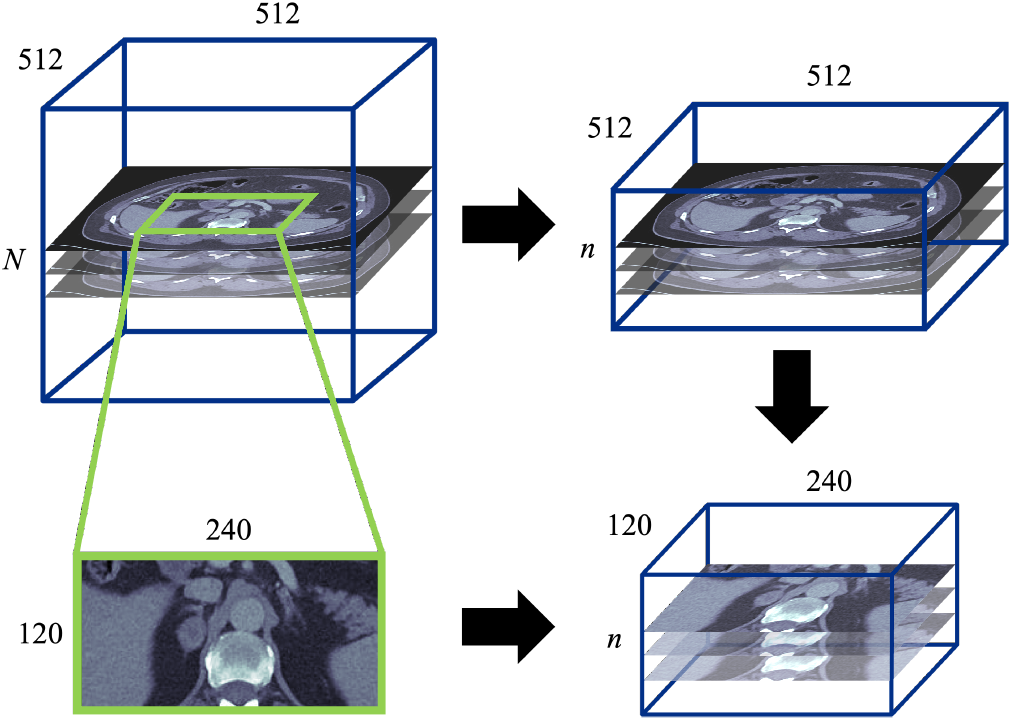
Illustration of the cropping process to focus on the ROI. (Top left) The original 3D CT scan with size of 512 × 512 × *N* pixel; (top right) a 1D cropping to select the *n* number of axial slices that covered the adrenals from the original scan; (bottom left) a 2D cropping that focus on the 240 × 120 pixel area on the axial slice that covered the adrenals; (bottom right) combining the cropping on all the three dimensions. The result of the cropped 3D image has a size of 240 × 120 × *n* pixel.

As shown in Figure 1 (top left panel), the original CT scans in the dataset have a resolution of 512 × 512 × *N* pixel, where *N* is the total number of slices of the original scan. The original axial slices covered the entire axial cross-section of the patients’ body. We selected a region of 240 × 120 pixel to cover both the left and right adrenals of the patients (Figure 1, bottom left panel). The cropping process also contained an axial slices selection to reduce the number of slices to *n* (Figure 1, top right panel), where *n* is the number of axial slices required to at least cover the entire adrenal glands on both sides. Within the 234 CT scans in the dataset, the value of *n* is ranged from 30 to 122 with a mean at ∼ 46 slides. Combining the two cropping processes, the final 3D images that focused on our ROI have a dimension of 240 ×120 ×*n* pixel (Figure 1, bottom right panel).

### 2.3 Training and test sets

Some patients in the dataset have records of more than one CT scan. To avoid leaking information to/from the independent test set during the model training process, the training and test sets are split in terms of patient instead of individual CT scan. We retained 20% of patients in the independent test set and the rest of 80% in the main training set (Table 1).

This resulted in 194 CT scans in the training set for model training and validation. The remaining 40 CT scans are in the independent test set which was kept separate until the final model performance test for this study (never involved in any model training and validating process). There are a total 8,340 axial slices included in the training set and 1,863 slices in the test set (Table 1).

### 3 2.5D deep learning model

We developed a 2.5D deep learning binary classification model to perform the adrenal lesion detection on 3D CT scans. The definition of 2.5D images, the preparation of 2.5D images from the 3D CT scans, the model architecture, and the model training process for our 2.5D model is discussed in this section. We also applied a traditional 3D deep learning model in our study for a comparison to the 2.5D model. The 3D model and its performance is discussed in brief in Appendix A.

### 3.1 2.5D images

A 2.5D image is a type of image that lies between a typical 2D and 3D image. It can retains some level of 3D features and can potentially be processed as a 2D image by deep learning models. A greyscale 2D image is two dimensional with a size of *x* × *y*, where *x* and *y* are the length and width of the 2D image. And for a greyscale 3D image (e.g., a CT scan), with a size of *x* × *y* × *n*, it can be considered as a combination of a stack of n number of greyscale 2D images. The size of a 2.5D image is *x* × *y* × 3, and it represents a stack of 3 greyscale 2D images.

Typically, an extra dimension of pixel information is required to record and display 2D colour images in electronic systems, such as the three RGB (red, green, and blue) colour channels. This increases the size of a 2D image to *x*×*y*×3, where the 3 represents the three RGB channels. Many commonly used families of 2D deep learning algorithms (e.g., VGG, ResNet, DenseNet, and EfficientNet: Simonyan and Zisserman, 2015; He et al., 2016; Huang et al., 2016; Tan and Le, 2019) have taken colour images into account and have the ability to process images with the extra three channels. Taking advantage of the fact that pixel volumes have the same size between 2D colour images and 2.5D images, converting our CT scans data to 2.5D images can allow us to apply 2D deep learning models on our images.

Figure 2 demonstrates the concept of preparing a set of 2.5D images from a 3D CT scan. To generate a 2.5D image, we simply assembled three consecutive axial slices of the CT scan (e.g., slice 1, 2 and 3). This allowed us to generate *n* number of 2.5D images from a CT scan which has *n* axial slices (from a 2.5D image containing slice 1, 2, and 3 to slice *n* −2, *n* −1, and *n*). This provided a total number of 8,340 2.5D images in the training set and 1,863 in the independent test set. The binary labels (abnormal and normal) of all the 2.5D images are assigned the same as the ground truth labels from their associate CT scans in the dataset.

**Figure 2:**
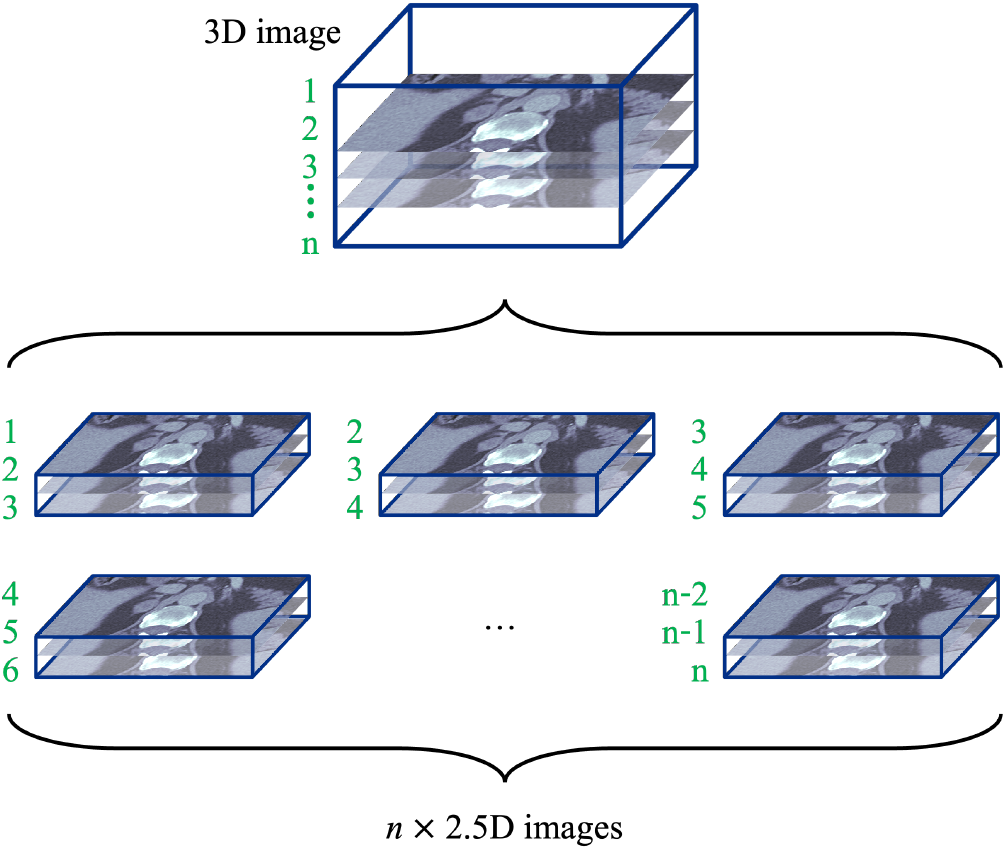
Illustration of the concept on generating a set of 2.5D images from a 3D CT scan (with *n* axial slices). The number coloured in green represent the axial slice number of the CT scan.

### 3.2 Model architecture and training process

The backbone architecture for our deep learning model is a 2D convolutional neural network (CNN). We use the EfficientNet family (Tan and Le, 2019) architecture for the images features extraction. The model version used in this study is EfficientNetB1. We applied transfer learning and used the pre-trained ImageNet (Russakovsky et al., 2015) weight of EfficientNetB1 to train the classification model on our training set.

Each 2.5D image in the training set was processed through an image augmentation layer before feeding into the EfficientNetB1 model. This image augmentation process contained a set of augmentation parameters, including a random rotation (between ±0.05π), random spatial zoom (zoomed in by a range of 20%), random coarse dropout (randomly dropping pixels with a probability between 0 and 0.1, e.g., DeVries and Taylor, 2017), and random contrast (with a random contrast factor, *f*_cont_, between 0.8 and 1.2). For each original pixel value, *x*_orig_, the new pixel value, *x*_new_, from the random contrast augmentation is calculated by

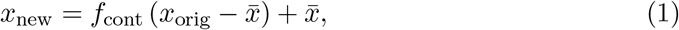

where 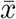 is the mean pixel value of the image slice (before augmentation).

The features extracted by EfficientNetB1 was then averaged out by a 2D global-average-pooling operation, and fed into a fully-connected layer (with random dropout rate of 0.2). The output layer of the model uses a softmax activation function to give the probabilities of the two classes (‘abnormal’ and ‘normal’).

The dataset (Table 1) shows a mild imbalance between the two classes in the training-validation set with a ratio of ∼ 1:1.8 abnormal-to-normal. This imbalance of classes should cause no significant deterioration to our model performance, however, we still includeed the class weighting in our training process.

We performed a 5-fold cross validation on our training set in the model training process. To prepare the 5-fold cross validation, we randomly split the patients in the training set into the five subsets. For each fold, we trained a model using one of these five subsets as the validation set and the other four subsets were combined into the training set (the training-validation ratio of 80% : 20%). Each subset is used once (but not more than once) as a validation set in the training process.

## 3.3 Model classification

### 3.3.1 Classifying 2.5D images

The classification of a 2.5D image is achieved by combining the five independent (fold-1 to fold-5) models trained during the 5-fold cross validation process. For each single independent model, the model prediction is calculated and provided in the form of classification probability of abnormal, *P*_abn_. We considered the 2.5D images as abnormal if the majority (⩾ 3 out of 5 independent models) of the 5-fold models has *P*_abn_ ⩾ 0.5, and classified as normal if the majority have *P*_abn_ < 0.5.

It is worth mentioning that having weak adrenal lesion signals (low value of *P*_abn_) on the majority of the 2.5D images does not necessarily mean that the CT scan and the patient are classified as normal. In a cropped CT scan that covers the whole two adrenal glands, the lesion(s) might only be visible only on few axial slices (therefore on few 2.5D images). A further adjustment of the 2.5D model is needed to provide a more comprehensive prediction in the unit of CT scans (Section 3.3.2).

### 3.3.2 Classifying 3D CT scans

To perform a binary classification in the unit of CT scans (instead of a single 2.5D image), all the classification results (from the 5-fold cross validation) of the 2.5D images that were generated from their associated CT scan are considered. To connect the classification prediction results from the 5-fold cross validation model and the CT scan, we introduced an operating value for our model to provide the final classification. The CT scans are classified as normal if the number of 2.5D images with *P*_abn_ ⩾ 0.5 is lower than the threshold operating value (in percentage). For example, if the operating value is defined to be *X*_ov_, a CT scan will be considered as normal if there are >*X*_ov_ of its 2.5D images classified as normal by the 5-fold model.

The operating value *X*_ov_ is obtained by comparing the 2.5D model prediction on the training set. Using the 2.5D model prediction on the training set and applying different *X*_ov_ (ranging from 0 to 100%), the overall accuracy for the CT scan prediction can be calculated. The final value of *X*_ov_ is chosen by the value that provides the best accuracy in the binary classification in the unit of CT scan. The best value of *X*_ov_ are then embedded to our model to give the final classification in the unit of CT scans.

## 4 Performance and results

Our independent test set contains 40 CT scans (Section 2.3 and Table 1) from patients that are excluded from the training set and the 5-fold cross validation process. There are 19 normal CT scans and 21 abnormal CT scans in this test set, which gives a prevalence of 0.525 on our test set^1^. In total, there are 1,863 2.5D images generated from the test set (Section 3.1), 805 of them are labelled as normal and 1,058 are labelled as abnormal. The 95% confidence interval (CI is used to represent the 95% confidence interval hereafter) are calculated using bootstrapping with 5,000 resamples.

For the classification in the unit of 2.5D images (Section 3.3.1), the prediction result of the 5-fold cross validation modal are shown in Figure 3. The AUC (area under curve) of the ROC (receiver operating characteristic) curve is 0.92 (CI: 0.91 − 0.93). The classification gives a sensitivity of 0.71 (CI: 0.68 − 0.74) and specificity of 0.92 (CI:0.90 − 0.94) when considering the P_abn_ ⩾ 0.5 as the abnormal threshold.

**Figure 3:**
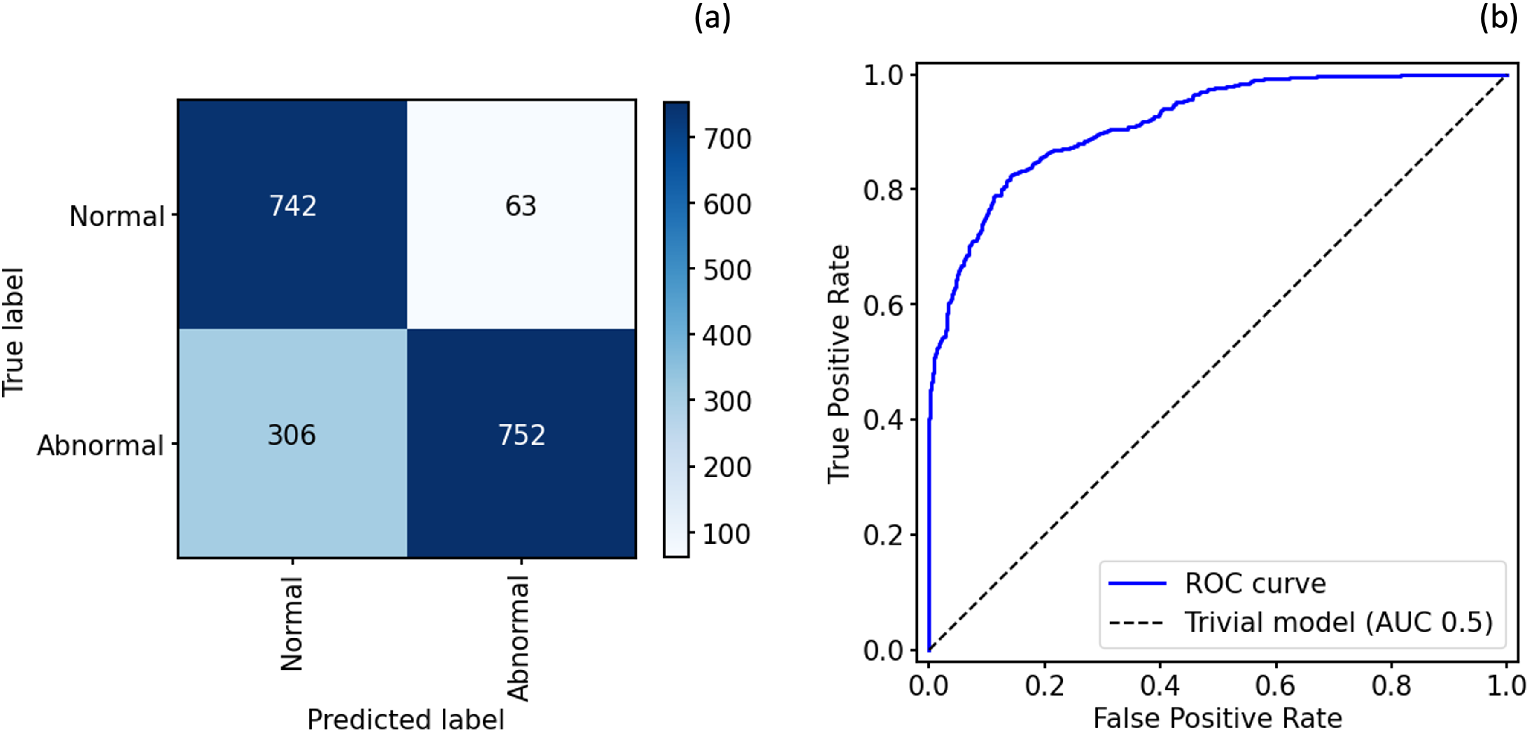
Performance of the 2.5D model on the independent test set. (a) The confusion matrix of the classification in the unit of 2.5D image. The true label is the ground-truth from the dataset, and the predicted label is the classification prediction from our 2.5D model. (b) The ROC curve of the 2.5D model. The AUC of the ROC curve is 0.92. The dashed line represents a trivial model (random guessing).

Figure 4 shows three classification examples in our test set by ‘naively’ setting *X*_ov_ = 50%. The example scan 1 and 2 (left and middle panels of Figure 4) show two correctly classified scans, where scan 1 (ground truth: normal) has almost all 2.5D images with *P*_abn_ < 0.5 and scan 2 (ground truth: abnormal) has a majority (> 50%) of 2.5D images with *P*_abn_ ⩾ 0.5. However, scan 3 (ground truth: abnormal) was wrongly classified as normal because most of the 2.5D images have *P*_abn_ < 0.5. As mentioned in Section 3.3.1, the adrenal lesion might only be covered by a few slices, setting a naive value of *X*_ov_ = 50% can lead to under-performance of the model.

**Figure 4:**
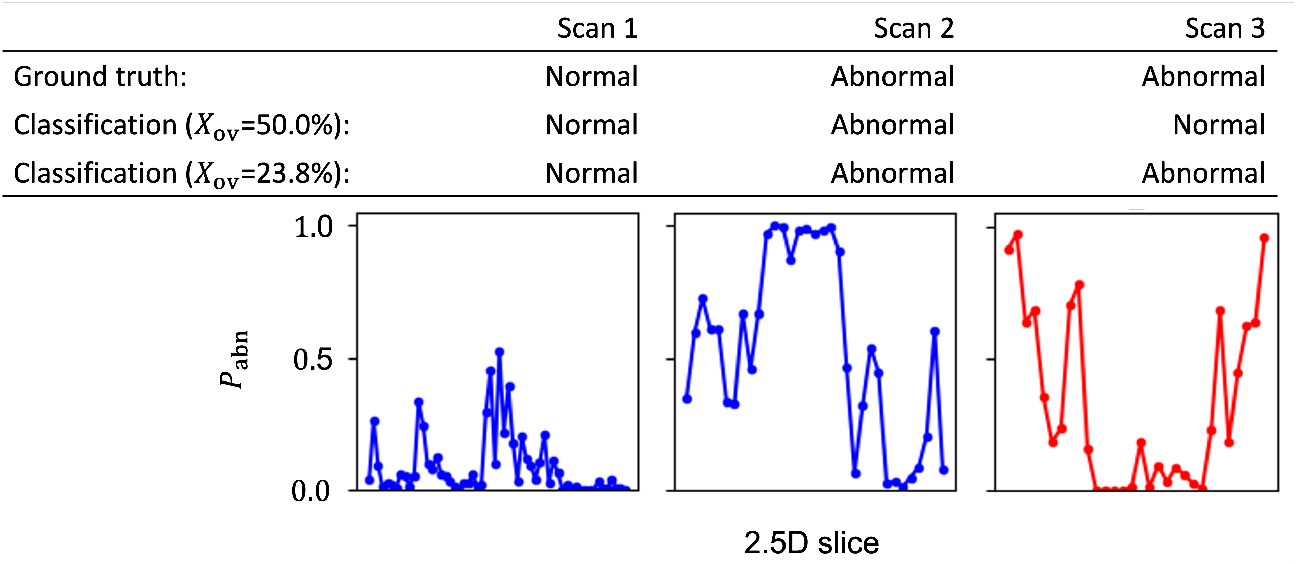
Three sample CT scans from the independent test set and the model prediction of them. The line plots show the prediction probability of being abnormal *P*_abn_ of the 2.5D images of the CT scans. Each data point (marked in dot) represents the value of *P*_abn_ for a 2.5D image. The sub-table on the upper panel listed the ground truth and model classification labels (with *X*_ov_ = 50.0% and 23.8%) of the three scans. The lines coloured in blue (scan 1 and 2) represent the classification model gives a correct classification (with both values of *X*_ov_). The line coloured in red (scan 3) represents the classification model gives at least one wrong classification (the wrong classification when adopting *X*_ov_ = 50.0%).

Applying the right value of *X*_ov_ can improve the model performance in classifying the 3D CT scans. The best operating value of *X*_ov_ for our model was computed by applying the 2.5D model on the training-validation set (Section 3.3.2). To avoid leaking data to the independent test set, this process of finding the best *X*_ov_ only obtained from the training-validation set and never calculated from the test set. Figure 5 shows the relative accuracy of the model prediction on the 3D scans with different *X*_ov_ (ranging from 0 to 100%). The relative accuracy peak at *X*_ov_ ≈ 23.8%. This represents if 23.8% of 2.5D images from the CT scan have *P*_abn_ ⩾ 0.5, that CT scan would be classified as abnormal by our model.

**Figure 5:**
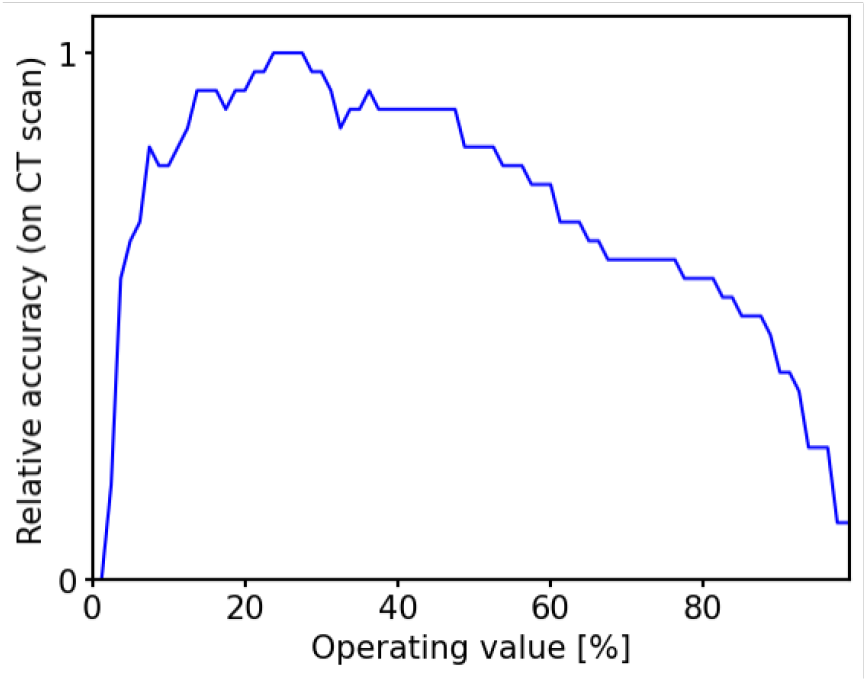
The variation between the operating value *X*_ov_ and the relative training-validation set accuracy of the classification in the unit of CT scans. The accuracy peaks (relative accuracy = 1) at *X*_ov_ = 23.8%.

Figure 6 shows the final 2.5D model classification performance (in the unit of CT scans). The value of *X*_ov_ = 23.8% is adopted in the model to perform this set of classification. The AUC of the ROC curve is 0.95. The classification gives a sensitivity of 0.86 and specificity of 0.89. The positive predictive value (PPV) has a value of 0.9 while the negative predictive value (NPV) has a value of 0.85.

**Figure 6:**
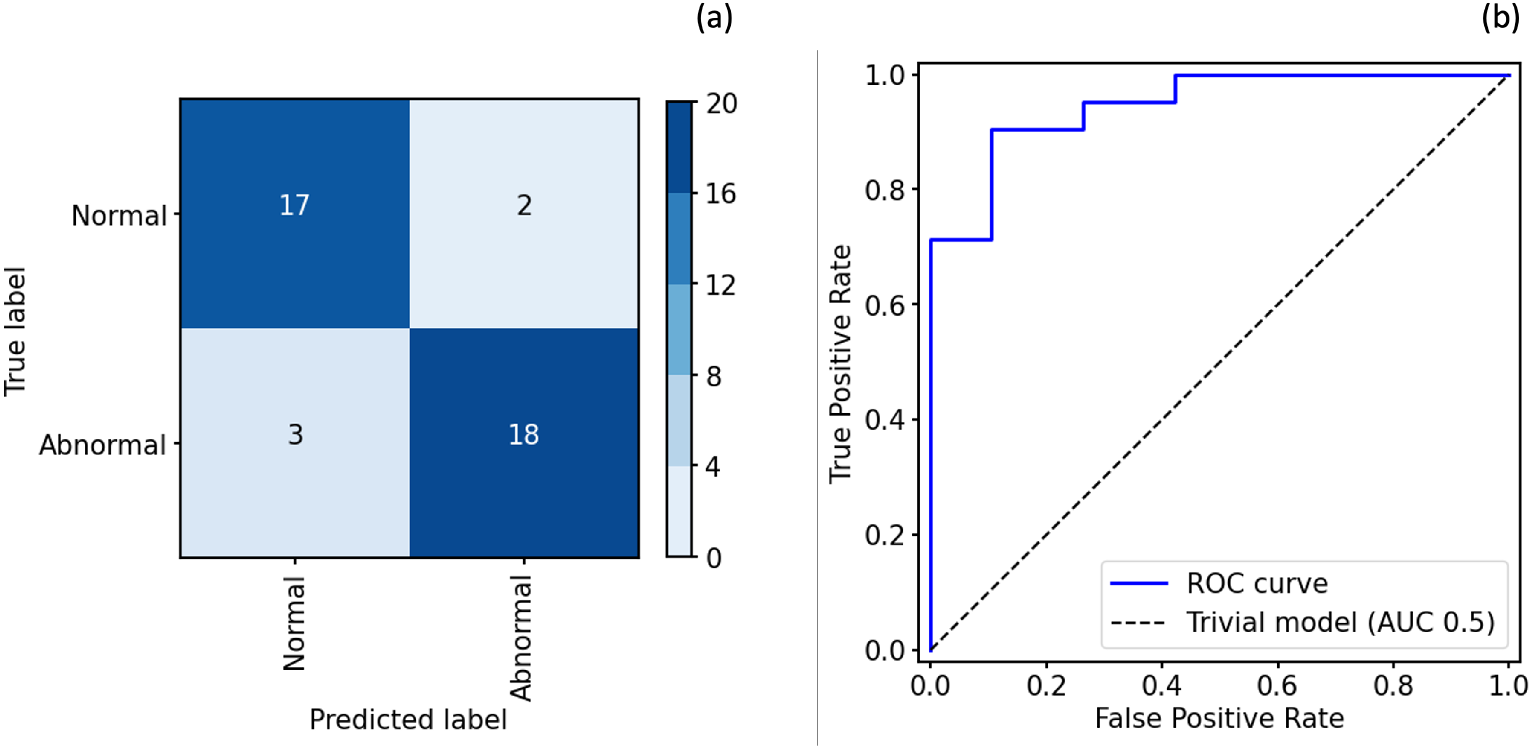
Performance of the 2.5D model on the independent test set (similar to Figure 3 but in the unit of CT scans instead of 2.5D images). (a) The confusion matrix of the 2.5D model (with *X*_ov_ = 23.8%) in the unit of 3D image. The true label is the ground-truth from the dataset, and the predicted label is the classification prediction on the CT scans from our 2.5D model. (b) The ROC curve of the 2.5D model on CT scans. The AUC of the ROC curve is 0.95. The dashed line represent a trivial model (random guessing).

## Discussion

We have presented the results of using deep learning models to detect adrenal lesions in CT scans. This examines whether or not whether or not adrenal lesions can be detected in CT scans by applying a 2.5D deep learning technique. Our approach to developing this proof-of-concept model was to train a deep learning model to perform image classification on patients’ CT scans, applying a 2.5D image transformation, to determine if the CT scans are capturing lesional (abnormal) or healthy (normal) adrenal glands.

The dataset used in this study contains 100 patients from the University Hospitals of North Midlands NHS Trust. The ratio between abnormal and normal patients is 1 : 1, and the ratio of abnormal and normal CT scans contained in the dataset is ∼ 1:1.6 (since multiple CT scans can be performed per patient). After applying the selection criteria (scans covering the whole adrenal glands, have a slice thickness between 0.5 and 3.0 mm, and have the same iodinated contrast density), there are 234 CT scans available in our dataset. We retained 20% of patients (20 patients with 40 CT scans in total) in our independent test set, which was only used for reporting the model performance in this paper (completely unseen for the model training, validation, and the process of calculating the operating value). Furthermore, we manually cropped the selected CT scans to our region of interest (adrenal glands on both sides of the patient). This 3D cropping gave a volume of 240 × 120 × *n* pixel that covered the two adrenal glands in all three dimensions.

To prepare the images for our 2.5D deep learning approach, we preprocessed all the selected CT scans further by transforming them to a set of 2.5D images. Each cropped CT scan with *n* axial slices can generate *n* number of 2.5D images, and each 2.5D image has a volume size of 240 × 120 × 3 pixels. In total, there are 8,340 2.5D images in our training set and 1,863 in the independent test set. Technically, the 2.5D images generated are a type of 3D images but only have an image depth (axial slices in this case) of 3. The three slices contained in a 2.5D image are three consecutive slices of their CT scan, where some 3D features are retained.

We developed our 2.5D deep learning model using EfficientNetB1 as our base architecture. Although EfficientNetB1 is a 2D CNN, it has the ability to process our 2.5D images because the CNN itself is designed for processing images with an image depth = 3 (commonly it applies on the three channels for the RGB colour of 2D images). During the training phase, we performed a 5-fold cross validation process on the training set data. A set of image augmentation processes was also applied to the training samples on each fold.

Moreover, we also computed that the best operating value is at *X*_op_ = 23.8% from the training set. This threshold means that a CT scan is classified as abnormal if more than 23.8% of the 2.5D images are predicted to be abnormal (with *P*_abn_ ⩾ 0.5). The reason why this *X*_op_ threshold was applied is due to the nature of many adrenal lesions only appearing on a small number of axial slices within the whole CT scan. Naively setting *X*_op_ = 50% might not truly reflect the underlying performance of the model and lead to missing many abnormal scans (a high number of false negatives).

Applying the 5-fold model of 2.5D image classification on the independent test set gives the AUC of the ROC curve of 0.92, sensitivity of 0.71, and specificity of 0.92. Compared with the values of AUC and specificity, the value of sensitivity is relatively low. It is mainly caused by the high number of false negative predictions. One of the reasons that induced this high number of false negative is due to the labelling of the 2.5D images. In our dataset for this study, the only ground truth labels are the binary labels (abnormal and normal) in the unit of CT scans, and the locations of the adrenal lesions are completely unknown within the scans. As we assigned to the 2.5D images the same labels as given by the CT scan, and the adrenal lesion might not affect the whole adrenal glands (lesions visible only on a few axial slices within the cropped scans), many 2.5D images were labelled as abnormal but did not carry signals of adrenal lesions. This resulted in some 2.5D images that were taken from an abnormal CT scan actually being correctly predicted to be normal because they do not carry signals of adrenal lesion, and covered only a normal region of an abnormal patient. Despite this introducing some extra noise (due to the limitation of the dataset), the presented model and its performance still managed to demonstrate a satisfactory level of ability to classify between abnormal and normal in the unit of 2.5D images.

Adopting *X*_op_ = 23.8% (value calculated only from the training set and completely independent from the test set) on the final 3D CT scan classification can reduce the impact caused by the data noise mentioned above. This set a higher threshold for classifying a CT scan as abnormal (from the overall prediction of all its 2.5D images). In the unit of CT scans, our classification model yields a sensitivity = 0.86, specificity =0.89, PPV = 0.9, NPV = 0.85, and an AUC of the ROC curve of 0.95 on the independent test set.

Comparing to a traditional 3D CNN model (Appendix A), our 2.5D model gives a better performance when tested on the same data. One of the drawbacks of using traditional 3D CNN models in this study is the small number of samples in our dataset. The performance result presented in Appendix A suggested that training such 3D CNN models with only 194 3D image samples might be difficult for the model to achieve a good performance. Using a 2.5D model approach has several advantages which improve the model predictive performance in this study. One of the advantages is the increase of the number of data points for model training (from 194 CT scans to 8,340 2.5D images). The evaluation of the test performance can also benefit from an increase number of data points in the test set (from 40 CT scans to 1,863 2.5D images). Furthermore, the backbone of the 2.5D model is a 2D CNN, which is smaller in size and less demanding on computer resources than a 3D CNN. It also allows the model to access a wider knowledge base (ImageNet, Russakovsky et al., 2015) through transfer learning for the model training.

We performed a proof-of-concept study using a 2.5D deep learning classification model to detect adrenal lesions on 3D CT scans. The results and performance of our 2.5D deep learning model demonstrate the ability and potential of applying such deep learning techniques on detecting adrenal lesions on CT scans. It also shows an opportunity to detect adrenal incidentalomas using deep learning. Nonetheless, we are aware that the proof-of-concept study presented in this paper is subject to some limitations. A small dataset from a non-diversified source may work well for the proof-of-concept but developing more robust prediction models can be difficult with datasets of this size. Acquiring extra information about the CT scans (e.g., the location of the lesions) in the dataset can also help to improve the model training by reducing the data noise. In future work, we will consider training an automated cropping tool to focus on the region of interest (adrenal glands), augmenting the dataset (increase the number of samples and improve the diversity of the data sources) and apply different training processes to improve the model robustness (e.g., knowledge distillation, Hinton et al., 2015). Further follow-up study will be conducted to advance our current model beyond a proof-of-concept.

## Data Availability

Algorithm code base is published on GitHub. Data and the models weights for transfer learning underlying this article will be shared on reasonable request to the corresponding author.

https://github.com/nhsx/skunkworks-adrenal-lesions-detection

## Acknowledgements

The authors wish to thank Amadeus Stevenson and Oludare Akinlolu for their contributions to the project. This research was conducted by the NHS AI Lab Skunkworks, an NHS England programme, in partnership with University Hospitals of North Midlands NHS Trust. The project was funded by the NHS AI Lab Skunkworks and made use of imaging data provided by the University Hospitals of North Midlands NHS Trust for the sole use of this project.

## Data availability

Algorithm code base is published on GitHub.^2^ Data and the models weights for transfer learning underlying this article will be shared on reasonable request to the corresponding author.

## Appendix A 3D deep learning model

Alongside to the 2.5D model (Section 3), we also investigated the approach of applying a 3D CNN to our use case.

### A.1 Data and the 3D model

Following the same 3D images acquisition procedure described in Section 2, we have 234 3D CT images (cropped to focus on the ROI) available for the 3D model training and testing (same as the examples considered in the main body of this paper: 194 images in the training set and 40 images in the independent test set).

Throughout the dataset, the minimum number of axial slices required to cover the entire adrenal glands on both side, *n*_min_, is 30. To avoid any upsampling on the 3D images, we downsampled all the images in the dataset to a size of 240 × 120 × 30 pixel for the 3D model training and testing.

The base architecture of our 3D deep learning model is a 3D residual network model (3D ResNet). The 3D ResNet model adopted in this study follow the architecture suggested by Solovyev et al. (2022), which transformed to 3D based on the original 2D ResNet model family (He et al., 2016). For the model training and performance testing demonstrated in this appendix, the 3D ResNet34 was chosen as the kernel model architecture.

The features extracted by the 3D ResNet34 are then averaged out by a 3D global-average-pooling operation, and fed into a fully-connected layer (with random dropout rate of 0.4). The output layer of the model uses a softmax activation function to give the probabilities of the two classes (‘abnormal’ and ‘normal’).

For the model training, we retained 20% of patients in the training set to be our validation set. The model was then trained on the remaining 80% of patients in the training set and the classification performance on the validation set was monitored. The final model weights are the set of parameters that provide the smallest validation loss during this 3D model training.

### A.2 Classification performance

The performance of the 3D classification model were tested on the independent test set (same test set for the 2.5D model performance test used in Section 4). Our 3D model has a moderate overall performance on the test set, providing an AUC-ROC of 0.79 (Figure 7) and a 95% CI at 0.64 and 0.91. For the classification predictions, we consider a CT scan to be an abnormal case if the prediction of the abnormal probability *P*_abnormal_ ⩾ 0.5. This resulted in a sensitivity of 0.62 and specificity of 0.68 (see also the confusion matrix of this classification in Figure 7).

**Figure 7:**
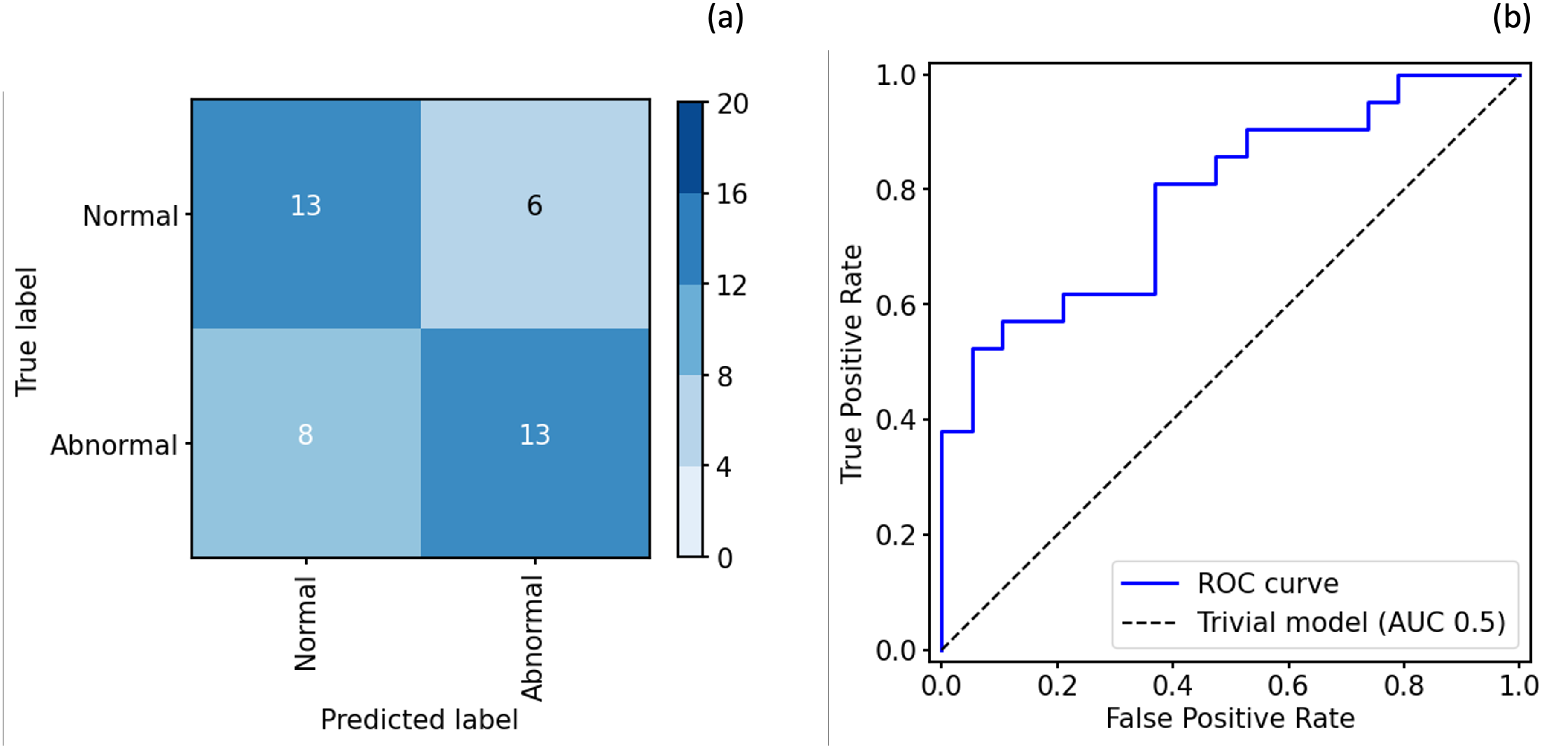
Similar to Figure 6 but for the performance of the 3D CNN model on the independent test set. (a) The confusion matrix of the 3D model. (b) the ROC curve of the 3D model. The AUC of the ROC curve is 0.79.

## Footnotes

1 The value of prevalence presented here represent only the distribution of our independent test set, and has no direct correlation to the prevalence of adrenal lesion in any wider population.

2 https://github.com/nhsx/skunkworks-adrenal-lesions-detection

## Notes

### Competing Interest Statement

The authors have declared no competing interest.

### Author Declarations

This proof of concept development project is not classified as research and therefore did not require ethical approvals. A stringent information governance process was followed, including fully anonymising patient data prior to sharing, and agreed by NHS England and the University Hospitals North Midlands NHS Trust information governance teams.

## References

M. Abecassls, M. McLoughlin, B. Langer, and J. Kudlow. Serendipitous adrenal masses: Prevalence, significance, and management. The American Journal of Surgery, 149(6): 783–788, 1985. ISSN 0002-9610. doi: https://doi.org/10.1016/S0002-9610(85)80186-0. URL https://www.sciencedirect.com/science/article/pii/S0002961085801860.

P. Anagnostis, A. Karagiannis, K. Tziomalos, A. I. Kakafika, V. G. Athyros, and D. P. Mikhailidis. Adrenal incidentaloma: a diagnostic challenge. Hormones, 8:163–184, 2009.

D. A. Bitter and D. S. Ross. Incidentally discovered adrenal masses. The American Journal of Surgery, 158(2):159–161, 1989. ISSN 0002-9610. doi: https://doi.org/10.1016/0002-9610(89)90367-X. URL https://www.sciencedirect.com/science/article/pii/000296108990367X. Papers of the Society for Clinical Vascular Surgery.

R. H. Caplan, P. J. Strutt, and G. G. Wickus. Subclinical Hormone Secretion by Incidentally Discovered Adrenal Masses. Archives of Surgery, 129(3):291–296, 03 1994. ISSN 0004-0010. doi: 10.1001/archsurg.1994.01420270067016. URL https://doi.org/10.1001/archsurg.1994.01420270067016.

T. DeVries and G. W. Taylor. Improved Regularization of Convolutional Neural Networks with Cutout. arXiv e-prints, art. arXiv:1708.04552, Aug. 2017.

M. Elmohr, D. Fuentes, M. Habra, P. Bhosale, A. Qayyum, E. Gates, A. Morshid, J. Hazle, and K. Elsayes. Machine learning-based texture analysis for differentiation of large adrenal cortical tumours on ct. Clinical Radiology, 74(10):818.e1–818.e7, 2019. ISSN 0009-9260. doi: https://doi.org/10.1016/j.crad.2019.06.021. URL https://www.sciencedirect.com/science/article/pii/S0009926019303162.

F. W. F. Hanna, B. G. Issa, S. C. Lea, C. George, A. Golash, M. Firn, S. Ogunmekan, E. Maddock, J. Sim, G. Xydopoulos, R. Fordham, and A. A. Fryer. Adrenal lesions found incidentally: how to improve clinical and cost-effectiveness. BMJ Open Quality, 9(1), 2020. doi: 10.1136/bmjoq-2018-000572. URL https://bmjopenquality.bmj.com/content/9/1/e000572.

K. He, X. Zhang, S. Ren, and J. Sun. Deep residual learning for image recognition. In 2016 IEEE Conference on Computer Vision and Pattern Recognition (CVPR), pages 770–778, Los Alamitos, CA, USA, jun 2016. IEEE Computer Society. doi: 10.1109/CVPR.2016.90. URL https://doi.ieeecomputersociety.org/10.1109/CVPR.2016.90.

G. Hinton, O. Vinyals, and J. Dean. Distilling the Knowledge in a Neural Network. arXiv e-prints, art. arXiv:1503.02531, Mar. 2015.

G. Huang, Z. Liu, L. van der Maaten, and K. Q. Weinberger. Densely Connected Convolutional Networks. arXiv e-prints, art. arXiv:1608.06993, Aug. 2016.

R. T. Kloos, M. D. Gross, I. R. Francis, M. Korobkin, and B. Shapiro. Incidentally Discovered Adrenal Masses*. Endocrine Reviews, 16(4):460–484, 08 1995. ISSN 0163-769X. doi: 10.1210/edrv-16-4-460. URL https://doi.org/10.1210/edrv-16-4-460.

K. Kruthika Rajeswari, and H. Maheshappa. Cbir system using capsule networks and 3d cnn for alzheimer’s disease diagnosis. Informatics in Medicine Unlocked, 14:59–68, 2019. ISSN 2352-9148. doi: https://doi.org/10.1016/j.imu.2018.12.001. URL https://www.sciencedirect.com/science/article/pii/S235291481830176X.

M. Kusunoki, T. Nakayama, A. Nishie, Y. Yamashita, K. Kikuchi, M. Eto, Y. Oda, and K. Ishigami. A deep learning-based approach for the diagnosis of adrenal adenoma: a new trial using ct. The British Journal of Radiology, 95(1135):20211066, 2022. doi: 10.1259/bjr.20211066. URL https://doi.org/10.1259/bjr.20211066. PMID: 35522787.

G. Mansmann, J. Lau, E. Balk, M. Rothberg, Y. Miyachi, and S. R. Bornstein. The Clinically Inapparent Adrenal Mass: Update in Diagnosis and Management. Endocrine Reviews, 25(2):309–340, 04 2004. ISSN 0163-769X. doi: 10.1210/er.2002-0031. URL https://doi.org/10.1210/er.2002-0031.

A. Moawad, A. Ahmed, D. Fuentes, J. Hazle, M. Habra, and K. Elsayes. Machine learning-based texture analysis for differentiation of radiologically indeterminate small adrenal tumors on adrenal protocol ct scans. Abdominal Radiology, 46, 10 2021. doi: 10.1007/s00261-021-03136-2.

D. Nie, H. Zhang, E. Adeli, L. Liu, and D. Shen. 3d deep learning for multi-modal imaging-guided survival time prediction of brain tumor patients. In S. Ourselin, L. Joskowicz, M. R. Sabuncu, G. Unal, and W. Wells, editors, Medical Image Computing and Computer-Assisted Intervention – MICCAI 2016, pages 212–220, Cham, 2016. Springer International Publishing. ISBN 978-3-319-46723-8.

O. Russakovsky, J. Deng, H. Su, J. Krause, S. Satheesh, S. Ma, Z. Huang, A. Karpathy, A. Khosla, M. Bernstein, A. C. Berg, and L. Fei-Fei. Imagenet large scale visual recognition challenge, 2015.

M. Sherlock, A. Scarsbrook, A. Abbas, S. Fraser, P. Limumpornpetch, R. Dineen, and P. M. Stewart. Adrenal Incidentaloma. Endocrine Reviews, 41(6):775–820, 04 2020. ISSN 0163-769X. doi: 10.1210/endrev/bnaa008. URL https://doi.org/10.1210/endrev/bnaa008.

K. Simonyan and A. Zisserman. Very deep convolutional networks for large-scale image recognition. 3rd International Conference on Learning Representations (ICLR 2015), pages 1–14, 2015.

S. P. Singh, L. Wang, S. Gupta, H. Goli, P. Padmanabhan, and B. Gulyás. 3d deep learning on medical images: A review. Sensors, 20(18), 2020. ISSN 1424-8220. doi: 10.3390/s20185097. URL https://www.mdpi.com/1424-8220/20/18/5097.

R. Solovyev, A. A. Kalinin, and T. Gabruseva. 3d convolutional neural networks for stalled brain capillary detection. Computers in Biology and Medicine, 141:105089, 2022. doi: 10.1016/j.compbiomed.2021.105089.

J. H. Song, F. S. Chaudhry, and W. W. Mayo-Smith. The incidental adrenal mass on ct: Prevalence of adrenal disease in 1,049 consecutive adrenal masses in patients with no known malignancy. American Journal of Roentgenology, 190(5):1163–1168, 2008. doi: 10.2214/AJR.07.2799. URL https://doi.org/10.2214/AJR.07.2799. PMID: 18430826.

M. Tan and Q. Le. EfficientNet: Rethinking model scaling for convolutional neural networks. In K. Chaudhuri and R. Salakhutdinov, editors, Proceedings of the 36th International Conference on Machine Learning, volume 97 of Proceedings of Machine Learning Research, pages 6105–6114. PMLR, 09–15 Jun 2019. URL https://proceedings.mlr.press/v97/tan19a.html.

X. Yi, X. Guan, C. Chen, Y. Zhang, Z. Zhang, M. Li, P. Liu, A. Yu, X. Long, L. Liu, B. T. Chen, and C. Zee. Adrenal incidentaloma: machine learning-based quantitative texture analysis of unenhanced ct can effectively differentiate spheo from lipid-poor adrenal adenoma. J Cancer, 9:3577–3582, 2018. doi: 10.7150/jca.26356. URL https://www.jcancer.org/v09p3577.htm.

J. Zhou, L.-Y. Luo, Q. Dou, H. Chen, C. Chen, G.-J. Li, Z.-F. Jiang, and P.-A. Heng. Weakly supervised 3d deep learning for breast cancer classification and localization of the lesions in mr images. Journal of Magnetic Resonance Imaging, 50(4):1144–1151, 2019. doi: https://doi.org/10.1002/jmri.26721. URL https://onlinelibrary.wiley.com/doi/abs/10.1002/jmri.26721.

